# Indications and Outcome of Intravitreal Bevacizumab Injection in a Community Eye Hospital, Nepal

**DOI:** 10.1101/2024.07.31.24311307

**Authors:** Sunil Thakali, Mohini Shrestha, Aleena Gauchan, Hom Bahadur Gurung, Manish Poudel

**Author notes:** First and Corresponding Author.

## Abstract

Intravitreal bevacizumab(IVB) injection, is a humanized monoclonal antibody that has been in use for the treatment of retinal diseases, very cheaply, especially for developing countries like Nepal. This is a retrospective study designed to evaluate the indications and outcomes of IVB at Hetauda Community Eye Hospital from 2019 to 2022. In this study, among 247 patients including 260 eyes with a follow-up rate of 221 patients involving 234 eyes, the mean patient age was 64.4 years, with male predominance of 56.1%. Thus, IVB was used principally in the treatment of diabetic retinopathy, neovascular age-related macular degeneration, and branch retinal vein occlusion. The results indicated significant improvements in central macular thickness and visual acuity with respect to diabetic retinopathy, nAMD, and BRVO. The study thus puts forth the effectiveness of IVB in improving visual outcomes and reducing CMT in a resource-constrained setting; hence, its use should be implemented as a viable treatment option within such an environment.

## Introduction

Bevacizumab is a monoclonal antibody against vascular endothelial growth factor –A(VEGF-A) that has been approved for metastatic colorectal carcinoma treatment since 2004. It is used off-label in various ocular diseases, it proved to be non-inferior to ranibizumab regarding both safety and efficacy[1, 2, 3].

Intravitreal Bevacizumab injections is used in the treatment of proliferative diabetic retinopathy, diabetic macular edema, choroidal neovascularization, retinopathy of prematurity, macular edema due to retinal vein occlusion, iris neovascularization, neovascular glaucoma, and pseudophakic macular edema[4, 5, 6].

Retinal diseases are significant cause for irreversible visual loss worldwide. A study in Bhaktapur, Nepal, showed that the prevalence of retinal diseases increased from 7.7% to 52.37% within four years among subjects aged 60 years and above. In Nepal, posterior segment diseases are the second major cause of blindness after cataract[7, 8].

IVB has shown to reduce VEGF levels in diabetic retinopathy and is less invasive than PRP, thus reducing the need for procedures such as pars plana vitrectomy[9, 10, 11].

Studies, like the BERVOLT study, and many meta-analyses have demonstrated that the patients with RVO significantly improved after IVB injections[12, 13].

Our study has several peculiarities: a single comprehensive ophthalmologist performed the IVB injections for all patients and represents a direct comparison with other centers and other specialties. Few studies have been published regarding indications and prevalence of IVB injections for retinal diseases in the Nepalese population [6, 14, 15] and the effect of IVB injection in DME [16, 17]. In addition, there has been no study of such kind in this part of Nepal. The primary aim of this study was to determine the indications and outcomes of intravitreal bevacizumab injections for different retinal diseases in this region.

## Material and Method

### Ethics statement

Ethical clearance was received from the Institutional review board of Tilganga Institute of Ophthalmology with reference number 18/2023 on 3^rd^ October 2023.

### Study Design

This was a retrospective study of retinal disease patients presenting to Hetauda Community Eye Hospital (HCEH) who have undergone Intravitreal Bevacizumab Injections(IVB) between 2019 to 2022. Data collection was started from 5^th^ November 2023 after ethical approval from Institutional review board of Tilganga Institute of Ophthalmology. In each visit the data of visual acuity on Snellen chart (converted to logarithm of minimum angle of resolution [log MAR]), intraocular pressure measurement on Goldman applanation tonometry, fundus examination, and central macular thickness measurement of the retina with SD-OCT (3D OCT-1 Maestro, Topcon) of each visits were taken from electronic medical record.

### Injection technique

All injections were performed in the operating room. Bevacizumab (Genentech Inc., San Francisco, CA, USA) 1.25 mg/0.05 mL, was aspirated into a 1 mL syringe with a 20/23 G needle from the vial (100mg/4mL). Each 1 ml syringe was capped with a 30 G needle ensuring no air in the syringe. Sterile gowns, caps, and masks were worn by the surgeon and patient. Eyelashes, eyelids, caruncle swabbed with Betadine 10%. Lid speculum and drape exposed the surgical area. They were injected at superotemporal using a 30-G needle at 3.5 or 4 mm posterior to the limbus in pseudophakia or phakic respectively with the patients looking down. Finally, ciprofloxacin ointment patching was done on the eye for two hours’ post-injection and treated with Ciprofloxacin eye drops four times daily for one week.

Written post-operative instructions were given to the patients and they were informed about the warning signs like ocular pain, decreased vision and lid edema. Follow up was done after 1 week, then 1 month, and subsequently every month. Reinjection was done after a minimum of four weeks from the date of primary injection. These are routine practices in Hetauda Community Eye Hospital.

### Data analysis

Data entry, cleaning, and coding were performed in Microsoft Excel and analyzed using IBM SPSS V.26. The Shapiro-Wilk test assessed the normality of pre-post vision and CMT data differences. Due to non-normal distributions, the Wilcoxon signed-rank test was used, with a p-value < 0.05 considered significant.

CMT improvement was calculated as baseline CMT (cmt0) minus final CMT (cmtF), yielding a positive value if improved, zero if unchanged, and negative if worsened. Vision improvement was calculated as baseline log MAR vision (va0) minus final log MAR vision (vaF), similarly yielding a positive value if improved, zero if unchanged, and negative if worsened.

## Results

This study included a review of 418 injections given to 247 patients (260 eyes). Excluding 26 patients lost to follow-up, there were 234 eyes of 221 patients. Of the latter, 29 had no OCT as only visual acuity was analyzed for them. The mean age of the 221 patients was 64.4 ± 12 years, with ages ranging from 20 to 88 years. The sex composition included 124 males (56.1%) and 97 females (43.9%). Most of the IVB injections were administered to patients from Makwanpur district, followed by Bara district. Diabetic retinopathy, neovascular age-related macular degeneration, branch retinal vein occlusion, and central retinal vein occlusion were the common indications for IVB injections. In detail, demographic features and indications are summarized in Tables 1 and 2.

**Table 1.** Demographic characteristics.

| Characteristics | n (%) |
| --- | --- |
| **Laterality** |  |
| Bilateral | 13 (5.9) |
| Unilateral | 208 (94.1) |
| **Ethnicity** |  |
| Hill Brahman | 87 (39.4) |
| Hill/Mountain Janajati | 57 (25.8) |
| Hill Chhetri | 35 (15.8) |
| Newar | 22 (10.0) |
| Hill Dalit | 14 (6.3) |
| Tarai/Madhesi other castes | 5 (2.3) |
| Others | 1 (0.5) |
| **Address** |  |
| Makwanpur | 209 (94.6) |
| Bara | 5 (2.3) |
| Sarlahi | 3 (1.4) |
| Rautahat | 3 (1.4) |
| Chitwan | 1 (0.5) |

**Table 2.** Indication of Intravitreal Bevacizumab.

| **Diagnosis** | **n (%)** |
| --- | --- |
| Diabetic Retinopathy | 84 (35.9) |
| nAMD | 68 (29.1) |
| BRVO | 57 (24.4) |
| CRVO | 9 (3.8) |
| Pseudophakic Cystoid Macular Edema | 6 (2.6) |
| CSR | 5 (2.1) |
| Circinate Retinopathy | 2 (0.9) |
| Retinal Vasculitis with Vitreous bleed | 2 (0.9) |
| Hypertensive Retinopathy with CME | 1 (0.4) |
| Total | 234 (100.0) |

**nAMD**, neovascular age related macular degeneration; **BRVO**, Branch retinal vein occlusion, **CRVO**, Central retinal vein occlusion; **CSR**, Central serous chorioretinopathy; **CME**, Cystoid macular edema

The median baseline CMT was 366 μm, ranging from 72 to 824 μm, while the median baseline VA was 0.78 log MAR, ranging from 0 to 3.7 log MAR. The median CMT significantly improved to 236 μm (range 74-877 μm) at the end of follow-up (p < 0.001), while the median best-corrected VA remained 0.78 log MAR (p = 0.108). CMT showed a significant improvement in BRVO, while VA demonstrated a significant improvement in BRVO, nAMD, and DR patients. The detailed changes in CMT and VA across different pathologies are presented in Tables 3 and 4.

**Table 3.**
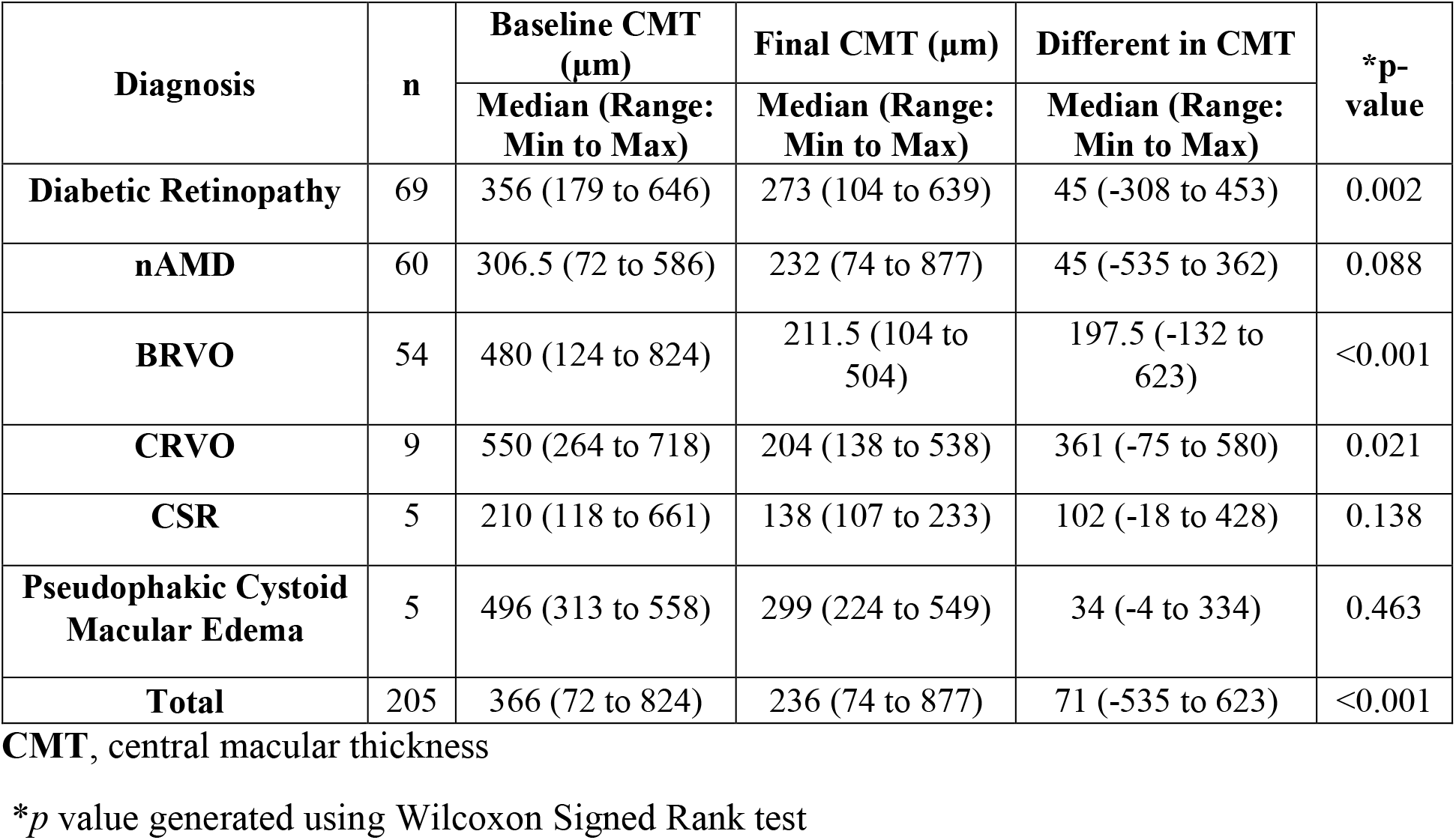
CMT at baseline and final.

| Diagnosis | n | Baseline CMT (μm) | Final CMT (μm) | Different in CMT | *p-value |
| --- | --- | --- | --- | --- | --- |
|  |  | Median (Range: Min to Max) | Median (Range: Min to Max) | Median (Range: Min to Max) |  |
| **Diabetic Retinopathy** | 69 | 356 (179 to 646) | 273 (104 to 639) | 45 (-308 to 453) | 0.002 |
| **nAMD** | 60 | 306.5 (72 to 586) | 232 (74 to 877) | 45 (-535 to 362) | 0.088 |
| **BRVO** | 54 | 480 (124 to 824) | 211.5 (104 to 504) | 197.5 (-132 to 623) | <0.001 |
| **CRVO** | 9 | 550 (264 to 718) | 204 (138 to 538) | 361 (-75 to 580) | 0.021 |
| **CSR** | 5 | 210 (118 to 661) | 138 (107 to 233) | 102 (-18 to 428) | 0.138 |
| **Pseudophakic Cystoid Macular Edema** | 5 | 496 (313 to 558) | 299 (224 to 549) | 34 (-4 to 334) | 0.463 |
| **Total** | 205 | 366 (72 to 824) | 236 (74 to 877) | 71 (-535 to 623) | <0.001 |

CMT, central macular thickness
\**p* value generated using Wilcoxon Signed Rank test

**Table 4.** VA at baseline and final.

| Diagnosis | n | Baseline VA | Final VA | Different in VA | *p-value |
| --- | --- | --- | --- | --- | --- |
|  |  | Median (Range: Min to Max) | Median (Range: Min to Max) | Median (Range: Min to Max) |  |
| **Diabetic Retinopathy** | 84 | 0.78 (0 to 3.7) | 0.6 (0 to 3.7) | 0 (-3.7 to 3.22) | <0.001 |
| **nAMD** | 68 | 1 (0.18 to 2.9) | 1 (0.18 to 2.9) | 0 (-2.42 to 2.42) | 0.001 |
| **BRVO** | 57 | 0.78 (0 to 2.9) | 0.6 (0 to 3.7) | 0.18 (-0.8 to 1.48) | <0.001 |
| **CRVO** | 9 | 1 (0.78 to 3.7) | 1 (0.3 to 3.7) | 0.22 (-0.3 to 1) | 0.021 |
| **Pseudophakic Cystoid Macular Edema** | 6 | 1 (0.6 to 1.78) | 0.78 (0.48 to 1.78) | 0.06 (-0.78 to 1) | 0.08 |
| **CSR** | 5 | 0.48 (0 to 1.78) | 0.48 (0 to 1) | 0 (-0.3 to 1.3) | 0.138 |
| **Total** | 234 | 0.78 (0 to 3.7) | 0.78 (0 to 3.7) | 0 (-3.7 to 3.7) | 0.108 |

**nAMD**, neovascular age related macular degeneration; **BRVO**, Branch retinal vein occlusion; **CRVO**, Central retinal vein occlusion; **CSR**, Central serous chorioretinopathy; **CME**, Cystoid macular edema; **VA**, visual acuity; **log MAR**, logarithm of the minimum angle of resolution.
\**p* value generated using Wilcoxon Signed Rank test

The number of IVB injections for various retinal diseases is shown in Table 5. DR has 84 injections in total, which makes up 35.9% of all injections. nAMD follows with 68 injections, which are 29.1%. BRVO is the third most prevalent with 57 injections, or 24.4%. The number of patients decreases significantly with the increasing number of injections.

**Table 5.**
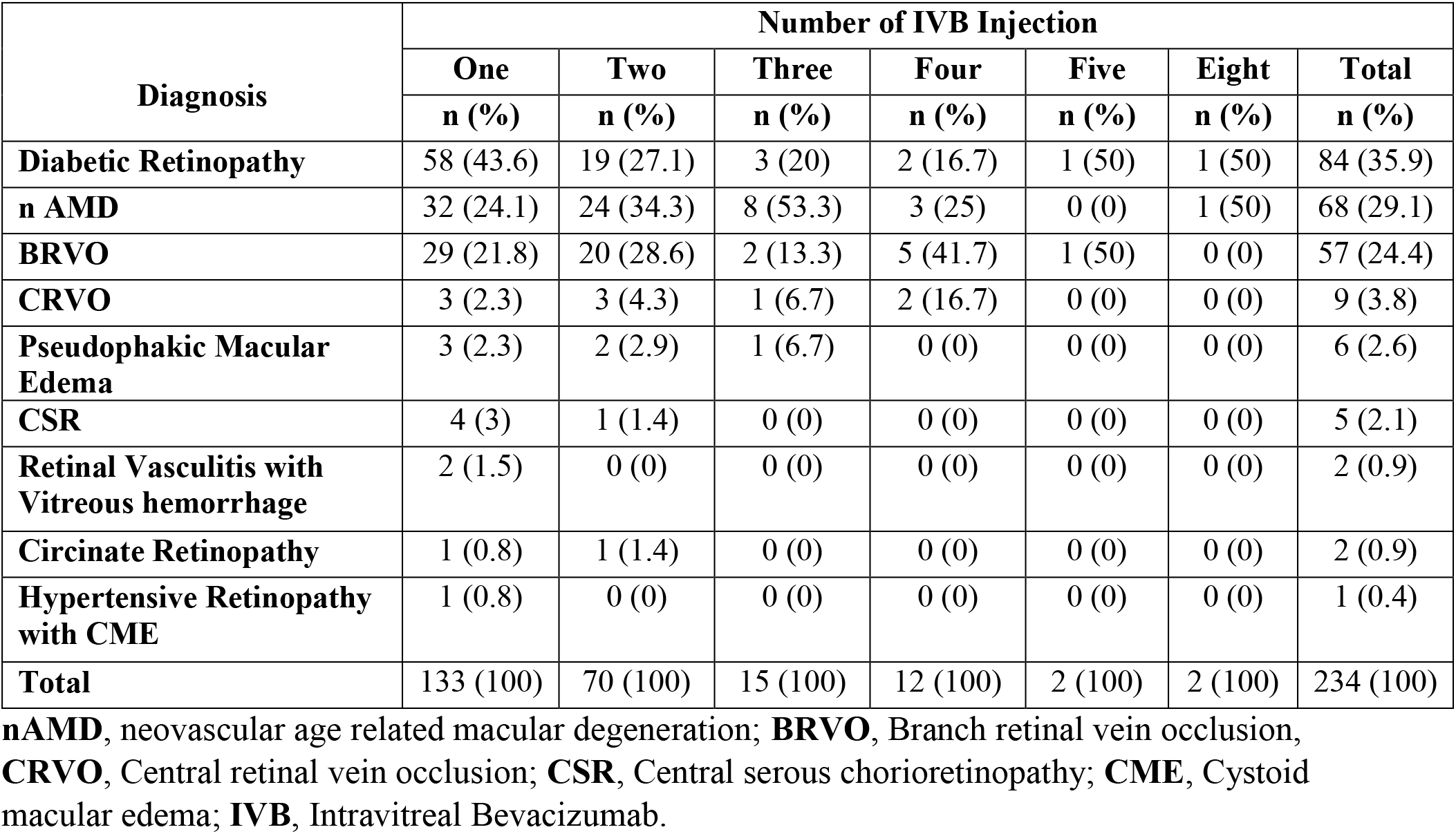
The number of IVB injections to various retinal diseases.

| Diagnosis | Number of IVB Injection |  |  |  |  |  |  |
| --- | --- | --- | --- | --- | --- | --- | --- |
|  | One | Two | Three | Four | Five | Eight | Total |
|  | n (%) | n (%) | n (%) | n (%) | n (%) | n (%) | n (%) |
| **Diabetic Retinopathy** | 58 (43.6) | 19 (27.1) | 3 (20) | 2 (16.7) | 1 (50) | 1 (50) | 84 (35.9) |
| **n AMD** | 32 (24.1) | 24 (34.3) | 8 (53.3) | 3 (25) | 0 (0) | 1 (50) | 68 (29.1) |
| **BRVO** | 29 (21.8) | 20 (28.6) | 2 (13.3) | 5 (41.7) | 1 (50) | 0 (0) | 57 (24.4) |
| **CRVO** | 3 (2.3) | 3 (4.3) | 1 (6.7) | 2 (16.7) | 0 (0) | 0 (0) | 9 (3.8) |
| **Pseudophakic Macular Edema** | 3 (2.3) | 2 (2.9) | 1 (6.7) | 0 (0) | 0 (0) | 0 (0) | 6 (2.6) |
| **CSR** | 4 (3) | 1 (1.4) | 0 (0) | 0 (0) | 0 (0) | 0 (0) | 5 (2.1) |
| **Retinal Vasculitis with Vitreous hemorrhage** | 2 (1.5) | 0 (0) | 0 (0) | 0 (0) | 0 (0) | 0 (0) | 2 (0.9) |
| **Circinate Retinopathy** | 1 (0.8) | 1 (1.4) | 0 (0) | 0 (0) | 0 (0) | 0 (0) | 2 (0.9) |
| **Hypertensive Retinopathy with CME** | 1 (0.8) | 0 (0) | 0 (0) | 0 (0) | 0 (0) | 0 (0) | 1 (0.4) |
| **Total** | 133 (100) | 70 (100) | 15 (100) | 12 (100) | 2 (100) | 2 (100) | 234 (100) |

**nAMD**, neovascular age related macular degeneration; **BRVO**, Branch retinal vein occlusion, **CRVO**, Central retinal vein occlusion; **CSR**, Central serous chorioretinopathy; **CME**, Cystoid macular edema; **IVB**, Intravitreal Bevacizumab.

## Discussion

The mean age of 221 patients in our study was 64.4 years ± 12 years. The minimum age being 20 years and the maximum age 88 years. It was similar to the study done at Kathmandu where the mean age of the patients was 59.62 years with ages ranging from 19 to 91 years [14].

In our study, the most predominant indication for IVB injections was DR. This reflect the high prevalence of this disease and visual impairment attributed to diabetes mellitus. Indeed, pathologies like macular edema, vitreous hemorrhage, and tractional retinal detachment are major causes of blindness among the working population and blind millions worldwide[18, 19].The fact that DR was the major primary indication for IVB injections in our study concurs with studies conducted in Nepal, Uganda, and Pakistan and continues to specify its widespread impact[14, 15, 20, 21].

In our study, nAMD was the second most common indication for IVB injections, unlike others where it usually occupies the third position after DR and RVO [6, 14, 15, 21]. Increased in life expectancy of Nepali population in current situation have more risk for nAMD and cost-effectiveness of IVB injection have driven its widespread use, despite lacking formal licensing for ocular conditions. [6, 22, 23].

RVO, including both BRVO and CRVO, was the third most common indication. Some studies report higher IVB usage for RVO compared to nAMD, highlighting regional differences in treatment patterns and disease prevalence [14, 15, 20, 21, 24].

In our study, PCME was the fourth most common indication for IVB injection. Recent large trials have demonstrated that IVB injections improve macular thickness and visual acuity in PCME.[5]. CSR was the fifth indication for IVB in our study, correlating with findings from other studies in Nepal and Pakistan, where CSR was also reported as the least common indication for IVB use [14, 21].

IVB injection in circinate retinopathy is undertaken to reduce macular edema and decrease progression of retinal complication and improve visual outcome[25]. IVB injection was shown to decrease macular edema and improve vision in patients who had hypertensive retinopathy. The results support the potential value of intravitreal bevacizumab as a useful treatment option for this condition[26, 27]. In retinal vasculitis with vitreous bleeding, the use of intravitreal bevacizumab has effectively demonstrated activity in halting the progression to severe complications and definitive visual loss [28].

The effect of IVB injection was further examined by disease diagnosis, and the results showed a modest change in CMT, there was significant improvement of VA by IVB among patients suffering from diabetic retinopathy, which is corroborated by findings from a similar study [20]. The good effect of IVB in DR, for both CMT and VA, is reported in many previous studies [16, 17, 29, 30].

There was an improvement in CMT and VA for patients with BRVO, consistent with reports from other studies [24, 31, 32]. Our study showed that CMT and VA in patients with CRVO were not significantly changed. This contrasts with the studies that have shown significant improvements after IVB injection, being comparable to the improvements achieved with intravitreal injections of Ranibizumab [13]. Poor visual outcome in our study may be influenced by the undetected ischemic CRVO and variable timing of Bevacizumab injections. [33, 34]. Visual and anatomic outcome in our setting are inferior to other trials and studies may be due to the lack of implementation of optimal diagnostic and retreatment criteria.

In comparison, patients with RVO showed less VA improvement in IVB injection compared to those with nAMD [24]. The greater VA gain and CMT reduction in nAMD justify the off-label use of bevacizumab as a highly cost-effective intervention for this condition [22].

While in our study, VA and CMT did not show any significant change with the use of IVB for Pseudophakic cystoid macular edema, the Pan-American Collaborative Retina Study Group did report significant improvements[5]. Similarly, IVB in CSR showed no significant changes in our study, but it remains a promising treatment requiring further investigation[35].

Our study evaluated the effect of IVB injection for a variety of retinal diseases one month after injection. We found that, in general, IVB injection led to anatomical and functional improvements. Unlike most studies, which are focused on the condition, such as diabetic macular edema or retinal vein occlusion, our research considered a wider spectrum of retinal pathologies.

## Conclusion

As the population ages and the prevalence of retinal diseases increases, the need for effective treatments like IVB injections rises. Our study emphasizes the effectiveness of IVB in improving VA and reducing CMT across various retinal conditions, with the most common indications being DR, nAMD, BRVO, and CRVO. These findings would therefore support the continued use of IVB as a cost-effective and accessible treatment option for retinal diseases in regions with limited healthcare resources.

## Strength and Limitation of Study

IVB injections by comprehensive ophthalmologists in the community eye hospital have a tremendous influence on reducing visual morbidity due to retinal diseases like DR and nAMD. The strengths of this approach include accessibility, expertise, cost-effectiveness, and attaining and maintaining improved visual outcomes. It is also evidence for advocacy for access to treatment in such resource-limited settings. However, it was a retrospective study done with a shorter follow up.

## Data Availability

The data supporting the findings of this study are available from Hetauda Community Eye Hospital. Due to privacy and confidentiality concerns, access to the data is restricted. Researchers interested in accessing the data should contact the corresponding author for further information and to discuss potential data sharing arrangements

## Acknowledgement

The authors would like to thank Hetauda Community Eye Hospital for providing the opportunity to conduct this study. We also extend our gratitude to the patients and hospital staff for their cooperation throughout the research.

## Author Contributions

**Conceptualization:** Sunil Thakali, Hom Bahadur Gurung

**Formal analysis:** Sunil Thakali, Manish Poudel

**Investigation:** Mohini Shrestha, Aleena Gauchan

**Supervision:** Hom Bahadur Gurung, Manish Poudel

**Writing – original draft:** Sunil Thakali

**Writing – review & editing:** Mohini Shrestha, Hom Bahadur Gurung

